# Soluble CD27 as an indicator of autoimmune disease in severe psychiatric disorders

**DOI:** 10.64898/2026.04.16.26351038

**Authors:** Isa Lindqvist, Celien Tigchelaar, Annica J. Rasmusson, Mikaela Syk, Gunnel Nordmark, Aysegül Sakarya, Elisabeth Skoglund, Peter T Schmidt, Andreas Kindmark, Anthony R. Absalom, Anders Larsson, Joachim Burman, Janet L. Cunningham

**Author notes:** Corresponding author: Professor Janet L. Cunningham, Department of Medical Sciences, Clinical Psychiatry, Uppsala University, Uppsala, Sweden.

## Abstract

T-cell activation may contribute to the pathogenesis of severe psychiatric disorders. Soluble CD27 (sCD27) —a marker for T-cell activation and disease activity in several autoimmune disorders —was evaluated as a tool for distinguishing T-cell activity in patients with severe psychiatric disorders, multiple sclerosis (MS), and controls. We hypothesised that positive sCD27 (sCD27+) would be associated with comorbid autoimmune disease (AID).

sCD27 was measured in cerebrospinal fluid (CSF) and blood from a population enriched for suspected immunological comorbidity, the Immunopsychiatry Cohort (IP; n=115), and patients with MS (n=37) where levels in both groups were higher when compared with age matched controls undergoing surgery (n=154). CSF-sCD27+, defined as >97.5 percentile of controls, was confirmed as a sensitive marker for MS (AUC=0.941) as 88% were CSF-sCD27+ while only 22% were blood-sCD27+ (AUC=0.608). In IP, 23% were CSF-sCD27+ and 15% were blood-sCD27+. CSF-sCD27+ was associated with comorbid AID (X^2^=4.847, p =0.028;) and AID disease activity (OR=5.14, p=0.029) and associations with AID were stronger when CSF and/or blood sCD27+ were combined (X^2^=8.559, p=0.003). CSF-sCD27+ was also associated with pleocytosis, CSF-Total-tau, and CSF-NfL in IP. In patients with severe psychiatric disorders, the sCD27+ cases were more likely to have comorbid AID and established markers for neuroinflammation in CSF. Combining analyses of CSF and blood improved sensitivity and specificity for AID suggesting compartmentalized T-cell activation. Psychiatric symptoms may precede somatic symptoms or be the prominent symptom of AID and sCD27 is a candidate marker for identification of this subgroup.

## Background

Autoimmune disease (AID) may present primarily with psychiatric symptoms and there is an increased risk of receiving psychiatric diagnoses up to five years before an AID diagnosis (1). Moreover, novel forms of AID specifically related to psychiatric disorders are rapidly emerging – each with its own set of phenotypic characteristics that respond to common immunological treatment strategies (2-10). Objective markers for AID are not always present in early phases, so the diagnoses of AID are based on clinical evaluations of typical somatic phenotypes, other comorbid AID and family history. AIDs are managed by the respective medical specializations best suited to evaluate the targeted organ and symptoms (5, 7, 11). Psychiatric symptoms, although, described as early manifestations of many forms of AIDs are under-recognized in clinical settings (8, 12-20) and clinical findings are frequently inconclusive (1, 17, 21). A meta-analysis of randomized controlled trials with modern immunotherapies for AID reveal that they can alleviate psychiatric symptoms, even when the primary somatic disease shows limited response (22). Psychiatric symptoms are rarely the primary treatment target, and no consensus exist regarding the managements of psychiatric manifestations of AID when they dominate the clinical picture (23). Despite the fact that a substantial number of patients presenting with psychiatric manifestations of AID, psychiatry lacks standardized frameworks for diagnostic evaluation and treatment of AID.

CD27 belongs to the tumor necrosis factor (TNF) receptor superfamily (24) and is seen as a promising inflammatory biomarker secreted by activated T-cells (25). T-cell receptor/CD3 complex activation upregulates CD27 expression and induces release of its soluble form, sCD27 (26-28). The shedding of CD27 thereby reflects T-cell activation (29). Additionally, CD27 is a marker of memory B-cells and plays a role in B-cell activation essential for the expansion of antigen-activated T-cells (28, 30). In summary, an inflammatory state involving the adaptive immune system can cause up-regulation of CD27 expression and release of sCD27 (28).

CSF-sCD27, first suggested as a biomarker for MS in the 1990s, is now established as a marker for intrathecal T-cell-mediated inflammation (29, 31, 32). CSF-sCD27 levels correlate with relapse rates in Clinically isolated syndrome (CIS) (27), whereas plasma-sCD27 concentrations were shown not to differ between MS patients and healthy controls (26). Moreover, elevated CSF-sCD27 discriminates patients with autoimmune encephalitis, including anti-N-methyl-D-aspartate (NMDA)- and LGI1 encephalitis from symptomatic controls (25, 33, 34). In patients with Neuromyelitis Optica Spectrum Disorder, CD27^+^ B-cell levels are linked to disease activity and have been used to determine the number of Rituximab (RTX) infusions administered (35).

Plasma-sCD27 levels have shown to estimate AID activity both in Systemic Lupus Erythematosus and rheumatoid arthritis (36, 37). Plasma sCD27 was furthermore increased in Graves’ disease and distinguishes Graves’ disease from thyroid cancer (38). Together, these observations support the role of sCD27 in CSF and blood as a broad marker for both central forms of neuroinflammation across inflammatory neurological diseases (33, 39) and peripheral AID (36-38).

Novel markers for AID in patients with severe psychiatric disorders are needed to reduce diagnostic delays and enable timely implementation of precision treatments (2, 40). This study builds on our previous findings that CSF-sCD27 serves as a general marker of central neuroinflammation (33). The hypothesis for this study emerged from our exploratory analyses of longitudinal measurements of sCD27 over 2.5 years from three patients with chronic, treatment refractory Obsessive-Compulsive Disorder (OCD), where repeated RTX treatment resulted in sustained clinical improvement (2). Out of these three cases, two showed positive CSF-sCD27 while the third had positive plasma-sCD27, and reductions in sCD27 followed clinical improvement identifying it as a candidate diagnostic and prognostic marker (2). Our study also introduced the possibility that both central and peripheral sampling may be of importance (2). In patients with relapsing-remitting multiple sclerosis (RRMS), drug naïve patients had significantly elevated CSF-sCD27 concentrations—median 354 pg/mL—and declined markedly following autologous hematopoietic stem cell transplantation, reaching 143 pg/mL at one year and continuing to decrease thereafter (41). The progressive normalization in these treatment studies support sCD27 as a dynamic biomarker reflecting immune activity.

As a crucial step towards determining clinical utility, we have recently studied the impact of factors including age, comorbidity, BMI and smoking on sCD27 levels in a larger and extensively characterized surgical cohort of patients undergoing spinal anesthesia mainly prior to orthopedical surgery from The Anesthetic Biobank of Cerebrospinal Fluid (ABC) established by the Department of Anesthesiology at the University Medical Centre Groningen (39, 42). Factors predicting CSF-sCD27 in the models had modest effect sizes and included plasma sCD27, age, and albumin quotient. Plasma levels of sCD27 were predicted by CSF-sCD27, plasma creatinine, BMI, and smoking, but surprisingly, also by psychiatric disorders (which had a prevalence of 9% in this population) which surpassed even neurological, hematological, and AID in importance when combined in the models (39).

This study aims to investigate whether sCD27—a marker of both T-cell activation and AID—is elevated in patients with severe psychiatric disorders compared to controls. We hypothesize that measuring sCD27 levels in CSF and blood and applying reference cutoff values determined using the 97.5th percentile of control values may identify patients with underlying T-cell activation and AID in a wide range of psychiatric disorders and potentially serve as a marker for novel forms of AID presenting with psychiatric symptoms. Patients with MS were used as a positive control for central neuroinflammation. The specificity and sensitivity of the sCD27 cutoffs were assessed in MS. In IP, the prevalence of sCD27+ in CSF and blood, and the ability of these cutoffs to differentiate between psychiatric patients with and without known AID was tested. Finally, we examined associations between sCD27 and established clinical markers of neuroinflammation, psychiatric symptoms, diagnostic categories and clinical red flags for autoimmune psychiatric syndromes.

## Methods and materials

### Ethical considerations

The study conforms to the provisions of the Helsinki Declaration and received ethical approval from the Regional Ethical Review Board in Uppsala, Sweden, under the reference numbers Drs 2012/081; 2014/148; 2008/182; 2013/278; 2015/462 and 2023-07214-02. The use of biomaterial and data from patients enrolled in the Anaesthetic Biobank of Cerebrospinal Fluid (ABC) was approved by the Medical Ethical Committee of the University Medical Centre Groningen (UMCG), The Netherlands (registration number 2016–174). All participants provided written informed consent.

### Surgical Controls

The Anesthetic Biobank of Cerebrospinal Fluid (ABC) was established by the Department of Anesthesiology at the UMCG in 2016 (42). Participants were patients undergoing spinal anesthesia for elective surgery. Data on sCD27 in relation to demographics, comorbidity, medical history, medication use, from the entire cohort has recently been published (39). For this study, a subset of surgical patients were selected as controls n=158, based on previously determined criteria (39), primarily excluding those with ASA Physical status III and IV (patients with severe systemic disease); neurological, psychiatric or AID; severe cardiovascular or kidney disease; use of psychotropic medication; smoking more than five cigarettes daily; alcohol abuse (more than two units per day); recreational drug use; drinking or smoking habits unknown, current or past malignant tumor. See exclusion chart in Supplementary Figure 1.

### Neuroinflammation (MS) Controls

A cohort of 37 MS patients who underwent routine care and underwent CSF and serum sampling between 2014 and 2015 served as a positive control for neuroinflammation. CSF but not serum from these patients has been included in the previous study (33). Inclusion criteria for this study were: i) a neurological diagnosis of MS according to standard diagnostic criteria, ii) access to medical records and laboratory results made in routine health care, iii) paired CSF and/or blood samples were available.

### Immunopsychiatry cohort (IP)

Samples from IP were collected at Uppsala University Hospital in Sweden, utilizing data gathered between 2012 and 2020. IP comprises patients displaying severe psychiatric disorders with clinical indications of potential immunological involvement. This population overlaps with a recently described study cohort (43) (see exclusion chart in Supplementary Figure 1). Whenever available, longitudinal data on immunological comorbidity were included. Comorbid AID, and AID activity were identified from the medical records using the International Classification of Diseases (ICD) 10 list of AID and verified by specialists in rheumatology, neurology, gastroenterology and endocrinology.

### Psychiatric and neurological assessment

The clinical assessment involved psychiatric interview, neurological examination, psychometric testing, and a review of the patient’s medical records (for details see (43)). Briefly, obsessions and compulsions were documented when they caused distress and/or dysfunction. Psychosis was defined as the presence of hallucinations, delusions, disorganized thought processes or behavior. Cognitive dysfunction was characterized by recent memory loss, executive dysfunction, or disorientation. Catatonia was noted if reported by the assessing / treating clinician, or if the patient had a total Bush-Francis Catatonia Rating Scale (BFCRS) score of more than three. Other psychiatric symptoms, such as agitation, mania, affective symptoms (including depression, anxiety, affective lability), behavioral regression, psychomotor retardation, tics, sleep dysregulation, and anhedonia were also documented. A relapsing-remitting course of illness was defined if the patient experienced major symptom fluctuations or had been asymptomatic for more than 1 month after symptom onset. All neurological examinations were conducted within 3 months of the CSF collection, most often on the same day. The presence of clinical red flags for suspected autoimmune psychiatric disease was assessed retrospectively, based on previously proposed consensus criteria and red and yellow flags for possible autoimmune encephalitis, psychosis, or OCD (11, 44, 45) (see Supplementary Table 1).

Data concerning comorbid AID was gathered from patients at sampling, and thereafter longitudinal data from before and after the CSF sampling were gathered from the medical records of follow-up visits until spring 2024. A panel of blinded clinical specialists in rheumatology, neurology, endocrinology and gastroenterology were asked to assess cases with comorbid AID regarding the diagnostic validity, AID activity and severity in relation to time of sampling. AID activity at sampling was evaluated using disease specific standard measures as either subclinical/mild or moderate/severe based on symptoms and the clinical data available at the time of sampling. In cases where patients had more than one AID, the disease with highest activity at sampling was noted as the main diagnosis. No patients presented with equivalent levels of disease activity between coexisting AID. (See Table 2 for AID within IP).

### Blood and CSF sample collection and analysis

sCD27 for IP and MS was analyzed in CSF, collected under non-fasting conditions, centrifuged at 2400 x g for 7 minutes at room temperature and subsequently stored in 225 µL (micronic tubes) in - 80°C at the Uppsala Biobank within 4 hours of sampling. Routine CSF analyses, including IgG indices, age-related CSF/plasma albumin ratios (AQ), unmatched oligoclonal bands (OCBs), and white blood cell counts were conducted by an accredited medical laboratory. Detailed reference values are provided in the previous paper (43). CSF samples compromised by traumatic lumbar punctures were excluded from analysis. Measurements of CSF neurofilament light (NfL) and glial fibrillary acidic protein (GFAP) concentrations were performed using enzyme-linked immunoassays (ELISAs) as previously described (46, 47) at Sahlgrenska University Hospital. Total tau (t-Tau) concentration in the CSF was measured using Lumipulse technology according to the manufacturer’s instructions (Fujirebio, Ghent, Belgium), and the results were compared to age-dependent reference intervals (Supplementary Table 2) used clinically (43). Immunofluorescence assays were conducted to detect antibodies against the N-methyl-D-aspartate receptor (NMDAR) using transfected cells (Euroimmun, Lübeck, Germany).

### Quantification of sCD27

Analysis of sCD27 levels was done on all samples at the same time point by a commercial sandwich ELISA kit, (DY382, R&D Systems, Minneapolis, MN, USA) detailed in (33). A technician blinded to the clinical information performed the analysis according to the manufacturer’s instructions. In this experiment the total CV was approximately 6%.

All samples were analyzed with the same batch of reagents, and patient and control samples were randomly distributed on the microtiter plates. If the sCD27 concentrations exceeded the upper limit of detection or were below the lower limit of detection (LOD) of 50 pg/mL, the values were replaced with 401,000 pg/mL or 25 pg/mL (LOD/2), respectively. IP patients and controls were analysed using plasma samples, whereas serum samples were used for MS patients. Given the strong linear relationship between plasma and serum sCD27 levels (Pearson’s r =0.98, P=2.1 × 10^□□^), they were considered analytically comparable and referred to as blood (See Supplementary Figure 6).

### Statistics

Outliers in the control population were identified using Z-scores (Z > 3) and confirmed on visual inspection of the relevant plots (see Supplementary Figure 2). Normality of sCD27 was assessed using the Shapiro–Wilk test and histograms, and variance by Levene’s test; as sCD27 was not normally distributed, nonparametric tests were applied throughout. Confounder assessment was performed by evaluating associations between candidate covariates and both exposure and outcome variables. Cutoffs for sCD27 positivity in CSF and blood were set at the 97.5th percentile of age-stratified controls (<45 and ≥45 years). Group comparisons of sCD27 levels between cohorts were performed using the Mann–Whitney U test with continuity correction.

Group comparisons for binary categorical markers were performed using the chi-square test with Yates ‘continuity correction. When expected cell counts where low, Fisher’s exact test was applied. Post hoc pairwise comparisons were performed where appropriate.

Ordinal logistic regression was performed to evaluate the association between sCD27 positivity and AID activity, adjusting for age and sex. Model discrimination was assessed using receiver operating characteristic (ROC) curves and the area under the curve (AUC). Exploratory correlations between sCD27 and inflammatory markers were performed using Spearman’s method with FDR correction. All analyses were performed in R (v4.3.3). Detailed descriptions of pre-processing, data transformations, and visualization are provided in the Supplementary Methods.

## Results

### Cohort Characteristics

See Table 1 for a summary of the demographics for the cohorts. Initially, 44 cases were identified to have comorbid AID in the IP. One case was removed after reevaluation by specialists as the diagnosis was evaluated as non-autoimmune. Thirteen patients had more than one AID (see Table 2 of comorbid AID in the IP) Visual and statistical analysis (defined as Z-score (Z) >3) revealed outliers in the control population (n=4, one in CSF (Z=7.64); three in blood (Z=7.45, Z=5.89, Z=3.26)) which were removed from further analyses (see Supplementary Figure 1 and Supplementary Figure 2).

**Table 1.** Summary of the demographics of the cohorts.

| Cohorts | Controls | IP | MS |
| --- | --- | --- | --- |
| N (CSF:Blood) | 154 (122:154) | 115 (115:114) | 37 (34:37) |
| Age (Mean, IQR) | 46 (32-58) | 32 (21-38) | 39 (29-46) |
| Sex (F:M) | (81:73) | (56:59) | (23:14) |
| BMI (kg/m<sup>2</sup>) | 27 (23-30) | 24 (20-27) | - |
| Q-ALB>ref | Na | 14/114 | 7/37 |
| CSF-OCB>ref | Na | 9/106 | 32/37 |
| CSF cells >ref | 0/154 | 4/113 | 19/37 |
| IgG Index >ref | Na | 5/113 | 30/37 |
| NfL >ref | Na | 14/114 | 21/34 |
| CSF-sCD27 (median, range)<br>pg/ml | 136<br>(25-528) | 184<br>(25-3797) | 742<br>(101-2447) |
| CSF-sCD27+ | 3/122 | 26/115 (23%) | 30/34 (88%) |
| <45 years |  | 25/96 | 22/25 |
| ≥ 45 years |  | 9/19 | 8/9 |
| Blood-sCD27 (median, range) | 4286<br>(1851-21389) | 7801<br>(2153-401000) | 3204<br>(1167-330793) |
| Blood-sCD27+ | 3/154 | 17/114 (15%) | 8/37 (22%) |
| <45 years |  | 13/95 | 3/27 |
| ≥ 45 years |  | 4/19 | 5/10 |
| Diagnostic subgroup details <sup>1</sup> | Na | Psychosis (63) OCD<br>(48)<br>NDD (37)<br>Catatonia (10)<br>Anx/Aff (80) | RRMS (27) CIS (5)<br>PPMS (4)<br>SPMS (1) |

<sup>1</sup>6 patients with treatment (5 RRMS and 1 SPMS), others are treatment naïve.
Age, Sex or BMI was not significantly different between the three cohorts.
IP: Immunopsychiatry, MS: Multiple Sclerosis, CSF: Cerebrospinal fluid, IQR: Interquartile Range, BMI: Body Mass Index, Q-Alb: Albumin quotient, OCB: Unmatched oligoclonal bands, NfL: Neurofilament light protein, OCD: Obsessive-Compulsive Disorder, NDD: Neurodevelopmental disorders including Autism spectrum, Attention Deficit Disorder or Attention-Deficit/Hyperactivity Disorder or intellectual disability, Anx/Aff: Anxiety and/or Affective disorders, RRMS: Relapsing-Remitting Multiple Sclerosis, CIS: Clinically Isolated Syndrome, PPMS: Primary Progressive Multiple Sclerosis, SPMS: Secondary Progressive Multiple Sclerosis.

**Table 2.** Description of IP patients with comorbid autoimmune diagnoses (n=43)

| **Variable** | **n (%)** |
| --- | --- |
| **Demographic and biological characteristics** |  |
| Female sex | 26 (60.5%) |
| Median age (IQR) | 35 (27.5–44.5) |
| Any CNS Damage marker | 14 (32.6%) |
| CNS neuroinflammation | 11 (25.6%) |
| CSF-sCD27+ | 15 (34.9%) |
| Blood-sCD27+ | 9 (20.9%) |
| sCD27+ | 21 (48.8%) |
| **Type of Psychiatric Disorder** |  |
| Psychosis | 24 (55.8%) |
| OCD | 12 (27.9%) |
| NDD | 14 (32.6%) |
| Catatonia | 3 (7.0%) |
| Anx/Aff | 30 (69.8%) |
| Multiple psychiatric disorders | 34 (79.1%) |
| **Autoimmune Comorbidity n (% of patients with AID)** |  |
| Systemic lupus erythematosus | 10 (23.3%) |
| Autoimmune Thyroiditis | 6 (14.0%) |
| Inflammatory bowel disease | 6 (14.0%) |
| Type 1 Diabetes | 4 (9.3%) |
| Antiphospholipid syndrome | 3 (7.0%) |
| Celiac disease | 3 (7.0%) |
| Rheumatoid arthritis | 3 (7.0%) |
| Multiple Sclerosis | 2 (4.7%) |
| Psoriasis | 2 (4.7%) |
| Anterior uveitis | 1 (2.3%) |
| Kawasaki disease | 1 (2.3%) |
| Polymyalgia rheumatica | 1 (2.3%) |
| Multiple AID | 13 (30.2%) |
| IQR: Interquartile Range, CSF-sCD27+: Positive sCD27 in cerebrospinal fluid, Blood-sCD27+: Positive sCD27 in blood, sCD27+: Positive sCD27 in either cerebrospinal fluid or blood, OCD: Obsessive-compulsive disorder, AID: autoimmune disorders, Anx/Aff: Anxiety and/or Affective Symptoms.<br>Any CNS Damage marker includes: (t-Tau) Total-tau, (GFAP) Glial fibrillary acid protein and (NfL) Neurofilament light chain.<br>CNS neuroinflammation: Pleocytosis, IgG index, OCB: Unmatched oligoclonal bands, NDD: Neurodevelopmental disorders include: Autism spectrum attention deficit disorder, Attention-Deficit/Hyperactivity Disorder and/or intellectual disability. |  |

In 5% of the samples, CSF-sCD27 concentrations were below the lowest limit of detection (LOD) of 50 pg/mL and blood sCD27 levels exceeded the upper limit of quantification of 400,000 pg/mL in 1% of the samples. The median and interquartile ranges (IQRs) of sCD27 levels in IP patients, MS patients and controls are presented in Table 1. No patient had any signs of kidney disease so sCD27 levels were not corrected for creatinine values. Participants were stratified into two age groups for the analyses based on previous work (39).

sCD27 CSF levels were compared between individuals below 45 years of age (<45) and 45 years or above (≥ 45) (see Figure 1A-B). In the groupwise analyses for cases with age <45 years, CSF levels were significantly higher in both IP and MS compared to controls (p=4.2 e-05, p=1.8 e-10 respectively), while only IP were found significantly different from controls in blood (p=9.7 e-09). For patients with age ≥45, both IP (n=19) and MS ≥45 (n=10) sCD27 levels were significantly higher than controls for CSF (p=0.042, p=7.3 e-05) and blood (p=0.00011, p=0.029), respectively (See Figure 1A-B). This finding remained significant after controlling for sex, BMI and smoking. A weak, but statistically significant correlation was observed between sCD27 levels in CSF and blood (Spearman’s R =0.32, P=1.7 × 10 □ □) in IP. However, data visualization revealed minimal overlap between patients classified as positive in the two compartments. Notably, several patients with the highest sCD27 concentrations in CSF—including two found to have MS in the IP cohort—were classified as negative in serum (see Figure 1C-D).

**Figure 1.**
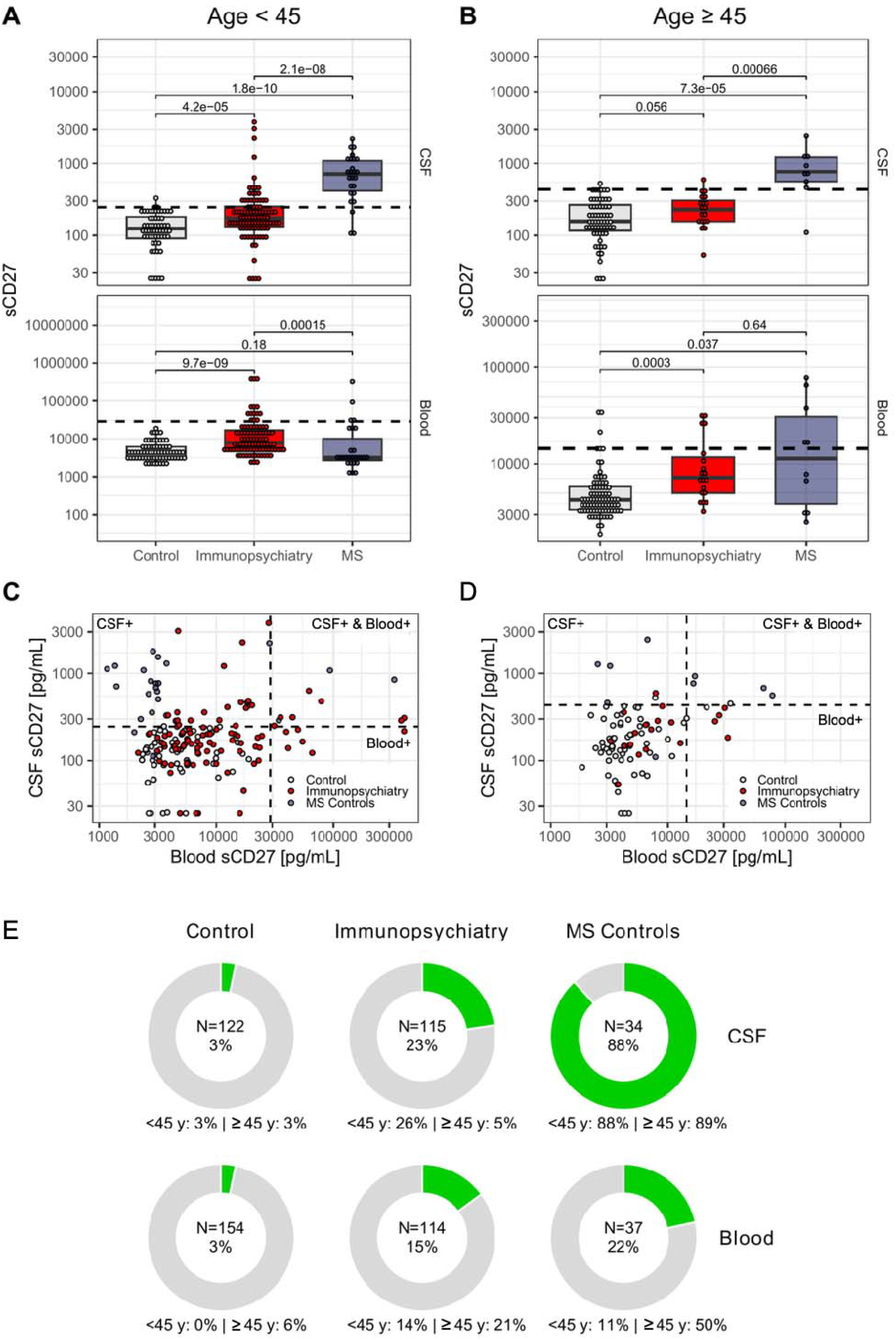
The distribution of sCD27 levels and positivity differed between cohorts. A-B) Groupwise comparisons of sCD27 levels (pg/ml) in CSF and blood in patients with severe psychiatric disorders (IP) and multiple sclerosis (MS) compared to controls < 45 (A) or ≥ 45 years old (B) (Mann Whitney U test). Black lines indicate 97.5 percentile of healthy controls sCD27 levels used to define positive cases. C-D) Scatter plots of CSF sCD27 vs blood sCD27 (pg/ml) in IP cohort shows that many patients in IP with the highest levels of sCD27 in CSF (CSF-sCD27+) do not exceed reference for positive values in blood and vice versa, suggesting compartmentalized immunological responses for < 45 (C) or ≥ 45 years old (D). Red dots represent the IP-cohort and white dots for healthy controls, the blue dots represent the MS subgroup. (E) Relative number of participants with sCD27 above reference values in the three cohorts. A log10 transformation is applied to all panels to modify only the spacing of the data points, achieving a more compressed y-axis that maintains proportional distances without altering the data’s original values on the axis labels.

Cutoff values for sCD27+ for the age groups <45 and ≥ 45 respectively, were 247 and 434 [pmol/ml] for CSF and 28927 and 14430 [pmol/mol] in blood, see Table 1 and Figure 1E.

### Clinical relevance, sCD27 and comorbid AID in IP

As shown in Figure 2A, CSF- and blood-sCD27+ was not confined to any specific psychiatric diagnostic category but were distributed across patients with diverse clinical manifestations (Table 3). Comorbid AID was present in 43 patients in IP where AIP was either known at sampling (n=34), was diagnosed within 16 months of sampling (n=7) or diagnosed since then (n=3). sCD27+ (blood and/or CSF) and CSF-sCD27+ was more likely in patients with AID compared to those without (X^2^=8.56, *p*=0.003 and X^2^=4.85, *p*=0.028) while blood-sCD27 values alone did not identify AID (X^2^=1.35, *p*=0.245). In CSF, sCD27 levels varied substantially across comorbid autoimmune disorders with the highest levels observed in patients with comorbid MS, NMDAr encephalitis or SLE (Supplementary Figure 4).

**Figure 2.**
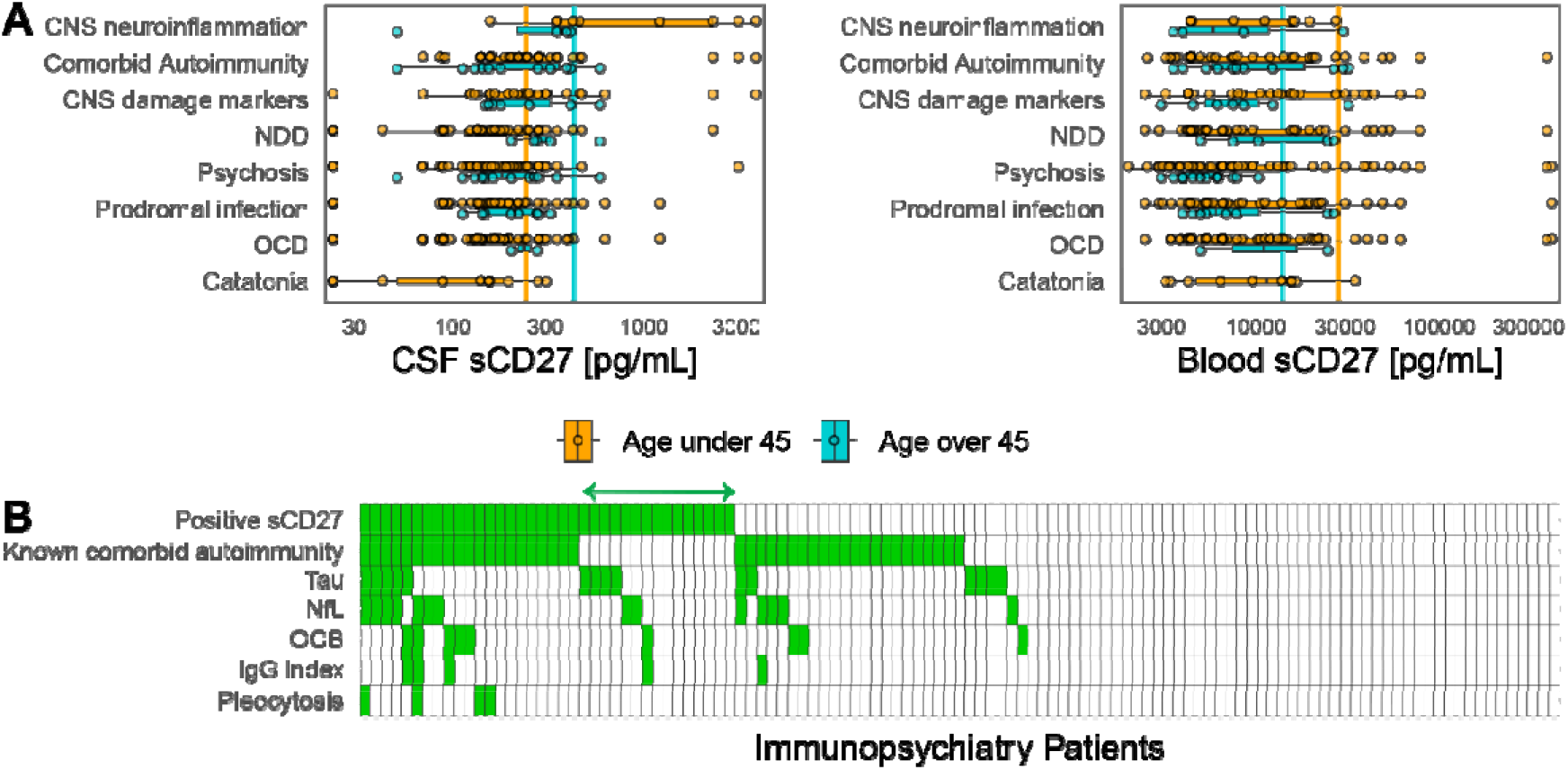
Distribution of sCD27 across clinical phenotypes and clinical markers in the Immunopsychiatry cohort (IP). A) sCD27+ in CSF and/or Blood was present in patients with different psychiatric disorders including psychosis, obsessive compulsive disorder (OCD), neurodevelopmental disorders (NDD; autism spectrum, attention deficit disorder, or Attention-Deficit/Hyperactivity Disorder, intellectual disability), and catatonia. CSF sCD27+ cases more frequently showed abnormal values for objective markers of central nervous system (CNS) pathology. Colours represent cases <45 years (orange) and cases ≥ 45 years (blue). Horizontal lines indicate the previously determined cutoff of the 97.5 percentile of controls for the respective age groups. Note that one individual may be presented more than once due to the possibility of having more than one trait simultaneously. B) The figure shows marker relate to elevated sCD27 levels where columns represent individual patients, and green indicates the presence of the clinical markers and known autoimmune disease. Green arrow in 3B shows sCD27+ in IP patients with no known autoimmune disorder and we suggest that T-cell activation may be contributing to the psychiatric phenotype. CNS neuroinflammation: Pleocytosis, IgG index, OCB: Unmatched oligoclonal bands. CNS damage markers: neurofilament light chain protein (NfL), total Tau protein (t-Tau) and glial fibrillary acidic protein (GFAP).

**Table 3.** Association between positive sCD27 levels and clinical markers in the Immunopsychiatry cohort.

|  | n (N) | sCD27+<br>n (CSF: B) | sCD27+<sup>1</sup> | X<sup>2</sup> | p-value<sup>2</sup> |
| --- | --- | --- | --- | --- | --- |
| **Assay** | 115 | 36 (26: 17) | 31.3% |  |  |
| IgG index | 5 (113) | 4 (4: 0) | 80% | 3.51 | 0.06 |
| CRP | 17 (98) | 6 (3:3) | 35% | 0.005 | 0.94 |
| CSF t-Tau | 15 (106) | 11 (8:3) | 73% | 4.78 | 0.03* |
| CSF NfL | 14 (114) | 11 (6:5) | 79% | 6.27 | 0.01* |
| CSF GFAP | 13 (112) | 7 (3:4) | 54% | 0 | 1 |
| CSF OCB | 9 (114) | 6 (5:1) | 67% | 3.51 | 0.06 |
| Pleocytosis<sup>3</sup> | 4 (113) | 4 (4:0) | 100% | - | 0.008* |
| Any abnormal CSF | 41 (106) | 21(17:9) | 51% | 8.72 | 0.003* |
| Comorbid AID | 43 (115) | 21 (15:9) | 49% | 8.56 | 0.003* |
| **Type of Psychiatric Disorder** |  |  |  |  |  |
| Psychosis | 63 (115) | 16(12:9) | 25% | 1.69 | 0.193 |
| OCD | 48 (115) | 15 (7:12) | 31% | 2.1 e<sup>-31</sup> | 1 |
| NDD | 37 (115) | 16 (11:9) | 43% | 2.84 | 0.092 |
| Catatonia<sup>3</sup> | 10 (115) | 2 (2:1) | 20% | 0.83 | 0.667 |
| Anx/Aff | 80 (115) | 24 (17:10) | 30% | 0.06 | 0.812 |

<sup>1</sup>Percentage of positive in both CSF-blood compartments
<sup>2</sup>The result is regarded significant if below 0.05 and marked with \*, Yate's continuity correction is used to get adjusted p-values.
<sup>3</sup>Fishers exact test was used due to the low frequency.
n: Number of positives, N: Total number analyzed, CSF: Cerebrospinal fluid, B: Blood, X<sup>2</sup>: Chi squared test, CRP: C-reactive protein, T-Tau: Total-Tau, NfL: Neurofilament light protein, GFAP: Glial Fibrillary Acidic protein, OCB: Unmatched oligoclonal bands, AID: Autoimmune disease, NDD: Neurodevelopmental disorders include: Autism spectrum attention deficit disorder, Attention-Deficit/Hyperactivity Disorder and/or intellectual disability, OCD: Obsessive-compulsive disorder, Anx/Aff =Anxiety and/or Affective disorders.

AID activity at sampling was rated as subclinical/mild in 20 patients and moderate/severe in 16 cases, whereas 7 cases could not be evaluated due to missing data. Both CD27+ (blood and/or CSF) (OR=4.81, *p*=0.038) and CSF-sCD27+ (OR=5.14, *p*=0.029) were related to moderate/severe AID when compared to subclinical/mild AID while, again, blood values alone were not related to activity (OR=0.43, *p*=0.355).

### Clinical symptoms and findings in patients with sCD27+ in IP

In the IP cohort, sCD27+ status was significantly associated with multiple markers of CSF pathology (Table 3). sCD27+ was more frequent in individuals with elevated CSF-t-Tau (X^2^=4.78, *p=*0.03) and increased CSF-NfL; (X^2^=6.27, *p=*0.01). Pleocytosis showed a strong association with sCD27+, with all individuals exhibiting pleocytosis being CSF-sCD27+ (*p=*0.008). When aggregating CSF abnormalities (t-Tau, NfL, GFAP, OCB, IgG Index and pleocytosis), individuals with any abnormal CSF marker were significantly more likely to be sCD27 positive compared with those without CSF abnormalities (X^2^=8.72, *p=*0.003).

The sCD27+ IP patients showed a wide range of psychiatric phenotypes and multiple psychiatric disorders (See Table 3), with no specific symptom or diagnosis predominating in univariate analyses. In IP, CSF-sCD27+ identified all patients with CSF pleocytosis (n=4) and comorbid AID was statistically more common (Table 3). Importantly, blood-sCD27 was negative in all four patients, indicating CNS derived sCD27. Of further interest, 15 of the IP patients were not diagnosed with any known comorbid AID and still had high sCD27 levels. Several of these patients had abnormal findings for other CSF markers (Figure 2B, green arrow).

EEG and MRI data was acquired in a subset of patients, but abnormalities were not associated with sCD27+ (data not shown).

To identify clinical psychiatric disease predictors for sCD27+, we employed feature importance ranking using random forest models based on clinical data (Supplementary Figure 5). Across CSF, blood, and combined CSF and blood data for sCD27+, the strongest predictors included, AID, acute symptom onset, presence of neurological symptoms, seizures, and neuroleptic malignant-syndrome. Comorbid AID was consistently among the top-ranking factors across all models.

To assess the ability of sCD27 cut-offs to discriminate between individuals with and without AID, we performed ROC analyses using logistic regression models, see Supplementary Figure 3. In the IP cohort, CSF sCD27 positivity showed a moderate discriminatory power (AUC=0.704 [0.605–0.790]) to predict comorbid AID, while blood sCD27 positivity alone yielded a lower performance (AUC=0.677 [0.593–0.759]). Combining blood and CSF sCD27+ showed best discrimination between autoimmune and non-autoimmune patients (AUC=0.751 [0.662–0.832]). This finding is consistent with the presence of both central, systemic and peripheral forms of AID in the cohort. Together, these analyses indicate that CSF-sCD27 in necessary for diagnostic accuracy but combining CSF and blood provides best accuracy for identifying both central and peripheral forms of autoimmune pathology.

### Clinical relevance, sCD27 and positive neurological CNS disease

To place the findings from the IP patients in context, corresponding analyses were performed in MS. The disease subtypes (CIS, RRMS, PPMS) and treatment effects in MS were not compared as the majority were classified as RRMS and were untreated. Subsequent ROC analyses using logistic regression models showed excellent discriminatory power between MS and controls for CSF-CD27+ (AUC=0.941 [0.883-0.986]) but not blood-sCD27+(AUC=0.608 [0.545-0.682]) (Supplementary Figure 3).

## Discussion

In line with our hypothesis, sCD27, a marker for T-cell activation, was elevated in CSF and/or blood in patients with severe psychiatric disorders compared with age-matched controls. The study moved beyond groupwise comparison to validate age stratified references for pathological levels of sCD27 in CSF and blood. Of particular importance, sCD27+ was compartment-specific and a subset of patients with showed isolated sCD27 elevation in CSF, with levels similar to patients with MS reflecting central nervous system but not peripheral involvement. Integrating CSF and blood measurements improved the identification of comorbid AID. Of particular importance. To our knowledge, this is the first study to systematically assess sCD27 across central and peripheral compartments in psychiatric disorders.

Notably, nearly a fourth of IP cohort showed CSF-sCD27 levels above reference and comparable to the MS cohort. Patients with MS served as a positive control for central neuroinflammation. Notably, our median CSF-sCD27 levels in psychiatric patients — when stratified by positivity — fall within the pre-treatment levels observed in RRMS (358 *vs* 354 pg/mL), suggesting a moderate but measurable degree of CNS immune activation in a subgroup within the IP cohort. This finding is of particular interest given that CSF-sCD27 showed high prediction in ROC curves for MS and that both MS and other central AID may initially manifest with psychiatric symptoms (1, 48). Importantly, patients with severe psychiatric disorders and sCD27+ were more likely to have or receive an AID and had higher clinical ratings of AID activity supporting the biological and clinical relevance of the applied thresholds and congruent with previous findings in CSF (27). The findings additionally indicated compartmentalised immunological activity. In the MS cohort as well as the two IP patients who were later found to have MS, sCD27 positivity was more prominent in CSF than in blood, reflecting CNS-compartmentalized pathology similar to previous findings (26, 29, 31, 32). In contrast, other patients in IP were identified with blood-sCD27+, high AID activity but CSF levels were below reference. This highlights the importance of conducting both CSF and blood sampling. In contrast to other studies focused on populations with SLE and RA (36, 37), blood-sCD27+ alone did not reach significant association with AID, as many patients with AID in IP were only sCD27+ in CSF. Blood-sCD27+ added value to identifying comorbid AID when added to those positive in CSF. We did not attempt to separate central from peripheral forms of AID in the classifications and our findings also raise questions about the traditional presumed localizations for inflammation. As an example, CSF-sCD27+ was observed in 3/5 patients with type 1 diabetes. This was an unexpected finding, as the condition is typically regarded as a peripheral and non-active, post-autoimmune disease, following initial beta-cell destruction, rather than a disease marked by ongoing immune activation (49). These findings implicate central T-cell activation in underlying mechanisms for associated psychiatric states that may be a consequence of the diabetes or possibly reflect yet to be diagnosed immunological comorbidities. Nearly one third of patients with AID had more than one cormorbid AID and it is also possible that underlying vulnerability factors may be independently contributing to psychiatric disorders.

Positive CSF-sCD27 was associated with known clinical markers for neuroinflammation, such as pleocytosis, and elevated NfL and T-Tau levels. A subgroup, 15 of the IP patients, were not diagnosed with any known comorbid AID were still sCD27+. Our case studied demonstrated that sCD27 distinguished patients without known comorbid AID who responded well to immunomodulatory RTX treatment (2). These observations imply that sCD27 levels above reference may help to identify underlying neuroinflammation or AID in severe psychiatric diseases, congruent with previous findings and the study hypotheses. Future work will aim to reveal if the sCD27+ precedes the emergence of a (somatic) autoimmune disorder or novel forms of psychiatric autoimmune disorders and evaluate implications for both psychiatric and immunological treatment responses. Specifically, we propose that sCD27 may be a marker that can reliably predict response to B-cell depletion therapy which would open new avenues for treatment in psychiatry (2, 11, 50).

Patients with sCD27+ presented with a wide range of psychiatric symptoms and most had received multiple psychiatric diagnoses. CSF-sCD27+ was particularly prominent among patients presenting with complex manifestations, including features such as disorganization, confusion, aggressiveness, affect dysregulation, concentration and memory impairment as well as olfactory and visual hallucinations. These symptoms are described for other states of active neuroinflammation and are in line with a role for sCD27 and T-cell–mediated processes in these patients.

Supporting the relevance of sCD27 as a candidate diagnostic and prognostic biomarker for immune-driven pathology in severe mental disorders , sCD27 has been shown to have a relatively short half-life, estimated to be <8 hours, with studies demonstrating rapid decline following for example surgical removal of malignant tissue (51). This transient kinetic profile implies that sustained elevations of sCD27 are unlikely to result from a past, resolved immune response, and instead point toward ongoing T-cell activation.

### Limitations

The age-stratified analyses may reduce statistical power compared with modelling age as a continuous variable. Age stratification reflects routine clinical practice, where biomarker interpretation is commonly based on predefined age-specific reference ranges rather than regression-based adjustments. Additionally, evaluating clinical relevance using binary classification metrics such as ROC curves and AUC may be limited, as these can overestimate discriminatory performance or mask clinically meaningful nuances (52). Although the control group was preselected to exclude comorbidities affecting the immune system the underlying surgical problem may still have influenced immunological activity, thereby impacting cutoff thresholds and prognostic predictions. This limitation is likely in a conservative direction, given the observed moderate specificity and low sensitivity. To partially mitigate this potential risk, four outliers were removed from the control population which resulted in lower cutoffs for positive results in the patient population. Controls with Z-scores between 2-3 were included and it is possible that the calculations might be too stringent, but at this early stage it is better to be conservative. The absence of positive controls for peripheral AID without psychiatric disease limited our ability to ascertain blood reference values’ sensitivity and specificity.

Elevated sCD27 levels may stem from underlying infections. However, patients showed no signs of infections and CRP levels were normal and sCD27 didn’t correlate with CRP in IP.

## Conclusion

These results provide clear support for sCD27, as a clinically relevant biomarker in psychiatric settings. This represents a crucial step toward bridging the gap between subjective clinical assessment and biological mechanisms in psychiatry. We validate previous findings that sCD27, when measured in both CSF and blood, is a reliable marker for neuroinflammation in cases with comorbid AID. We also suggest that sCD27 may be a potential marker for compartment-specific T-cell activation in patients with primarily psychiatric symptoms. In addition to the potential diagnostic utility, ours and others work indicate that sCD27 may also inform treatment decisions and prognosis. Further systematic studies are warranted, particularly to investigate the ability of sCD27 status to predict treatment response to immunomodulatory therapies in patients with psychiatric disorders.

## Data Availability

All data produced in the present study are available upon reasonable request to the authors

## Acknowledgements

The work in this study was supported by Åke Wiberg stiftelsen, Åhlens stiftelsen, Märta och Nicke Nasvells fund; The mental health Fund (Fonden för Psykisk Hälsa), Fredrik and Ingrid Thurings Stiftelse; Stiftelsen Söderström-Königska sjukhemmet; The Swedish Society of Medicine; ALF Funds from Uppsala University Hospital. The funding agencies had no influence in the study design, data collection, analysis, interpretation, writing, or publication decision. The authors wish to thank Svante Berg, Ida Deogun and Anna Branth for excellent clinical research assistance, and Uppsala Biobank for management of the samples. The authors also acknowledge Emma Tornvind’s contributions in examining the patients` medical histories. The authors are especially grateful to patients for providing medical records and samples to improve care.

## Author Contributions

Lindqvist, I, Rasmusson, A. J.: Methodology, formal analysis, data curation, visualization, writing– original draft, writing–review & editing. Tigchelaar, C., Syk, M., Nordmark, G., Skoglund, E., Schmidt, PT., Kindmark, A., Absalom, A. R., Larsson, A., Burman, J: Data acquisition, Writing– review & editing. Sakarya A.: Writing–review & editing. Cunningham, J. L: Conceptualization, methodology, formal analysis, writing–original draft writing–review & editing, funding acquisition; supervision.

## References

1. Marrie RA, Walld R, Bolton JM, Sareen J, Walker JR, Patten SB, et al. Rising incidence of psychiatric disorders before diagnosis of immune-mediated inflammatory disease. Epidemiology and psychiatric sciences. 2019;28(3):333–42.

2. Gallwitz M, Lindqvist I, Mulder J, Rasmusson AJ, Larsson A, Husén E, et al. Three cases with chronic obsessive compulsive disorder report gains in wellbeing and function following rituximab treatment. Molecular Psychiatry. 2024.

3. Kayser MS, Dalmau J. The Emerging Link Between Autoimmune Disorders and Neuropsychiatric Disease. The Journal of neuropsychiatry and clinical neurosciences. 2011;23(1):90–7.

4. Benros ME, Eaton WW, Mortensen PB. The epidemiologic evidence linking autoimmune diseases and psychosis. Biol Psychiatry. 2014;75(4):300–6.

5. Pollak TA, Lennox BR, Müller S, Benros ME, Prüss H, Tebartz van Elst L, et al. Autoimmune psychosis: an international consensus on an approach to the diagnosis and management of psychosis of suspected autoimmune origin. The Lancet Psychiatry. 2020;7(1):93–108.

6. Endres D, Pruss H, Rauer S, Suss P, Venhoff N, Feige B, et al. Probable Autoimmune Catatonia With Antibodies Against Cilia on Hippocampal Granule Cells and Highly Suspicious Cerebral FDG-Positron Emission Tomography Findings. Biol Psychiatry. 2020;87(9):e29–e31.

7. Endres D, Maier V, Leypoldt F, Wandinger K-P, Lennox B, Pollak TA, et al. Autoantibody-associated psychiatric syndromes: a systematic literature review resulting in 145 cases. Psychological Medicine. 2020;52(6):1135–46.

8. Isung J, Williams K, Isomura K, Gromark C, Hesselmark E, Lichtenstein P, et al. Association of Primary Humoral Immunodeficiencies With Psychiatric Disorders and Suicidal Behavior and the Role of Autoimmune Diseases. JAMA Psychiatry. 2020;77(11):1147–54.

9. Mulder J, Feresiadou A, Fällmar D, Frithiof R, Virhammar J, Rasmusson A, et al. Autoimmune Encephalitis Presenting With Malignant Catatonia in a 40-Year-Old Male Patient With COVID-19. The American journal of psychiatry. 2021;178(6):485–9.

10. Xu J, Liu RJ, Fahey S, Frick L, Leckman J, Vaccarino F, et al. Antibodies From Children With PANDAS Bind Specifically to Striatal Cholinergic Interneurons and Alter Their Activity. Am J Psychiatry. 2021;178(1):48–64.

11. Endres D, Pollak TA, Bechter K, Denzel D, Pitsch K, Nickel K, et al. Immunological causes of obsessive-compulsive disorder: is it time for the concept of an “autoimmune OCD” subtype? Transl Psychiatry. 2022;12(1):5.

12. Law C, Flaherty CV, Bandyopadhyay S. A Review of Psychiatric Comorbidity in Myasthenia Gravis. Cureus. 2020;12(7):e9184.

13. Vano-Galvan S, Egeberg A, Piraccini BM, Marwaha S, Reed C, Johansson E, et al. Characteristics and Management of Patients with Alopecia Areata and Selected Comorbid Conditions: Results from a Survey in Five European Countries. Dermatol Ther (Heidelb). 2024;14(4):1027–37.

14. Weiss DB, Dyrud J, House RM, Beresford TP. Psychiatric manifestations of autoimmune disorders. Curr Treat Options Neurol. 2005;7(5):413–7.

15. Etzioni A. Immune deficiency and autoimmunity. Autoimmun Rev. 2003;2(6):364–9.

16. Marrie RA, Walld R, Bolton JM, Sareen J, Walker JR, Patten SB, et al. Increased incidence of psychiatric disorders in immune-mediated inflammatory disease. J Psychosom Res. 2017;101:17–23.

17. Miller ZA, Sturm VE, Camsari GB, Karydas A, Yokoyama JS, Grinberg LT, et al. Increased prevalence of autoimmune disease within C9 and FTD/MND cohorts. Neurology, neuroimmunology & neuroinflammation. 2016;3(6):301.

18. Mataix-Cols D, Frans E, Pérez-Vigil A, Kuja-Halkola R, Gromark C, Isomura K, et al. A total-population multigenerational family clustering study of autoimmune diseases in obsessive-compulsive disorder and Tourette’s/chronic tic disorders. Mol Psychiatry. 2018;23(7):1652–8.

19. Ma M, Sandberg J, Farhadian B, Silverman M, Xie Y, Thienemann M, et al. Arthritis in Children with Psychiatric Deteriorations: A Case Series. Dev Neurosci. 2023;45(6):325–34.

20. Gerentes M, Pelissolo A, Rajagopal K, Tamouza R, Hamdani N. Obsessive-Compulsive Disorder: Autoimmunity and Neuroinflammation. Curr Psychiatry Rep. 2019;21(8):78.

21. Lu R, Munroe ME, Guthridge JM, Bean KM, Fife DA, Chen H, et al. Dysregulation of innate and adaptive serum mediators precedes systemic lupus erythematosus classification and improves prognostic accuracy of autoantibodies. Journal of auttoimmunity. 2016;74:182–93.

22. Wittenberg GM, Stylianou A, Zhang Y, Sun Y, Gupta A, Jagannatha PS, et al. Effects of immunomodulatory drugs on depressive symptoms: A mega-analysis of randomized, placebo-controlled clinical trials in inflammatory disorders. Mol Psychiatry. 2020;25(6):1275–85.

23. Cunningham JL, Nordmark G, Fällmar D, Cervenka S, Gallwitz M, Säll R, et al. Experiences in implementing immunopsychiatry in real life. Journal of Affective Disorders Reports. 2023;13:100597.

24. Han BK, Olsen NJ, Bottaro A. The CD27-CD70 pathway and pathogenesis of autoimmune disease. Semin Arthritis Rheum. 2016;45(4):496–501.

25. Wong YYM, Van Der Vuurst De Vries RM, Van Pelt ED, Ketelslegers IA, Melief M-J, Wierenga AF, et al. T-cell activation marker sCD27 is associated with clinically definite multiple sclerosis in childhood-acquired demyelinating syndromes. Multiple Sclerosis. 2018;24(13):1715–24.

26. Hintzen RQ, Van Lier RW, Kuijpers KC, Baars PA, Schaasberg W, Lucas CJ, et al. Elevated levels of a soluble form of the T cell activation antigen CD27 in cerebrospinal fluid of multiple sclerosis patients. Journal of neuroimmunology. 1991;35(1-3):211–7.

27. Van Der Vuurst De Vries RM, Mescheriakova JY, Runia TF, Jafari N, Siepman TAM, Hintzen RQ. Soluble CD27 Levels in Cerebrospinal Fluid as a Prognostic Biomarker in Clinically Isolated Syndrome. JAMA Neurology. 2017;74(3):286–92.

28. Hendriks J, Gravestein LA, Tesselaar K, Van Lier RAW, Schumacher TNM, Borst J. CD27 is required for generation and long-term maintenance of T cell immunity. Nature Immunology. 2000;1(5):433–40.

29. Cencioni MT, Magliozzi R, Palmisano I, Suwan K, Mensi A, Fuentes-Font L, et al. Soluble CD27 is an intrathecal biomarker of T-cell-mediated lesion. J Neuroinflammation. 2024;21(1):91.

30. Grimsholm O. CD27 on human memory B cells–more than just a surface marker. Clinical and experimental immunology. 2023;213(2):164–72.

31. Komori M, Blake A, Greenwood M, Lin YC, Kosa P, Ghazali D, et al. Cerebrospinal fluid markers reveal intrathecal inflammation in progressive multiple sclerosis. Annals of Neurology. 2015;78(1):3–20

32. Mahler MR, Søndergaard HB, Buhelt S, Von Essen MR, Romme Christensen J, Enevold C, et al. Multiplex assessment of cerebrospinal fluid biomarkers in multiple sclerosis. Multiple sclerosis and related disorders. 2020;45:102391.

33. Feresiadou A, Nilsson K, Ingelsson M, Press R, Kmezic I, Nygren I, et al. Measurement of sCD27 in the cerebrospinal fluid identifies patients with neuroinflammatory disease. Journal of Neuroimmunology. 2019;332:31–6.

34. Cobanovic S, Blaabjerg M, Illes Z, Nissen MS, Nielsen CH, Kondziella D, et al. Cerebrospinal fluid soluble CD27 is a sensitive biomarker of inflammation in autoimmune encephalitis. Journal of the Neurological Sciences 2024;466.

35. Bruschi N, Malentacchi M, Malucchi S, Sperli F, Martire S, Sala A, et al. Tailoring Rituximab According to CD27-Positive B-Cell versus CD19-Positive B-Cell Monitoring in Neuromyelitis Optica Spectrum Disorder and MOG-Associated Disease: Results from a Single-Center Study. Neurology and Therapy. 2023;12:1375–83.

36. Font J, Pallares L, Martorell J, Martinez E, Gaya A, Vives J, et al. Elevated Soluble CD27 Levels in Serum of Patients with Systemic Lupus Erythematosus. Clinical immunology and immunopathology. 1996;81(3):239–43.

37. Shi L, Yang J, Xu J, Dai J, Li J. Elevated serum soluble CD27 levels are associated with both disease activity and humoral immune activity in patients with rheumatoid arthritis. Clinical and experimental rheumatology. 2024;42(5).

38. Kallio P, Murphy ED. Soluble CD27 in thyroid disorders. J Lab Clin Med. 1998;132(6):478–82.

39. Tigchelaar C, Cunningham JL, Rasmusson AJ, Thulin M, Burman J, Kema IP, et al. Cerebrospinal fluid and plasma concentrations of the inflammatory marker soluble CD27 in a large surgical population. iScience. 2024;27(6).

40. Hughes HK, Yang H, Lesh TA, Carter CS, Ashwood P. Evidence of innate immune dysfunction in first-episode psychosis patients with accompanying mood disorder. Journal of Neuroinflammation. 2022;19:287.

41. Lundblad K, Zjukovskaja C, Larsson A, Cherif H, Kultima K, Burman J. CSF Concentrations of CXCL13 and sCD27 Before and After Autologous Hematopoietic Stem Cell Transplantation for Multiple Sclerosis. Neurology, Neuroimmunology & Neuroinflammation. 2023;10(5).

42. Tigchelaar C, Atmosoerodjo SD, van Faassen M, Wardenaar KJ, De Deyn PP, Schoevers RA, et al. The Anaesthetic Biobank of Cerebrospinal fluid: a unique repository for. Ann Transl Med. 2021;9(6):455 LID – 10.21037/atm–20–4498 [doi] LID – 455.

43. Syk M, Tornvind E, Gallwitz M, Fällmar D, Amandusson Å, Rothkegel H, et al. An exploratory study of the damage markers NfL, GFAP, and t-Tau, in cerebrospinal fluid and other findings from a patient cohort enriched for suspected autoimmune psychiatric disease. Translational psychiatry. 2024;14:304.

44. Pollak TA, Lennox BR, Muller S, Benros ME, Pruss H, Tebartz van Elst L, et al. Autoimmune psychosis: an international consensus on an approach to the diagnosis and management of psychosis of suspected autoimmune origin. Lancet Psychiatry. 2020;7(1):93–108.

45. Gole S, Anand A. Autoimmune Encephalitis2024. Available from: https://www.ncbi.nlm.nih.gov/books/NBK578203/.

46. Gaetani L, Höglund K, Parnetti L, Pujol-Calderon F, Becker B, Eusebi P, et al. A new enzyme-linked immunosorbent assay for neurofilament light in cerebrospinal fluid: analytical validation and clinical evaluation. Alzheimer’s research & therapy. 2018;10(1):8.

47. Rosengren LE, Ahlsén G, Belfrage M, Gillberg C, Haglid KG, Hamberger A. A sensitive ELISA for glial fibrillary acidic protein: application in CSF of children. Journal of Neuroscience Methods. 1992;44(2-3):113–9.

48. Maier HB, Stadler J, Deest-Gaubatz S, Borlak F, Turker SN, Konen FF, et al. The significance of cerebrospinal fluid analysis in the differential diagnosis of 564 psychiatric patients: Multiple sclerosis is more common than autoimmune-encephalitis. Psychiatry Res. 2024;333:115725.

49. Thompson PA-O, Pipella JA-O, Rutter GA-O, Gaisano HA-O, Santamaria PA-O. Islet autoimmunity in human type 1 diabetes: initiation and progression from the. Diabetologia. 2023;66(11):1971–82 LID – 10.007/s00125–023–5970–z [doi].

50. Bejerot S, Sigra Stein S, Welin E, Eklund D, Hylén U, Humble MB. Rituximab as an adjunctive treatment for schizophrenia spectrum disorder or obsessive-compulsive disorder: Two open-label pilot studies on treatment-resistant patients. Journal of Psychiatric Research. 2023;158:319–29.

51. Van Oers M, Pals S, Evers L, Van Der Schoot C, Koopman G, Bonfrer J, et al. Expression and release of CD27 in human B-cell malignancies. Blood. 1993;8(11):3430–6.

52. Muschelli J. ROC and AUC with a Binary Predictor: a Potentially Misleading Metric. Journal of classification. 2021;37(3):696–708.

