## Supplemental tables and figures for "Soluble CD27 as an indicator of autoimmune disease in severe psychiatric disorders"

### Supplementary

**Supplementary Figure 1. Exlusion Chart.**

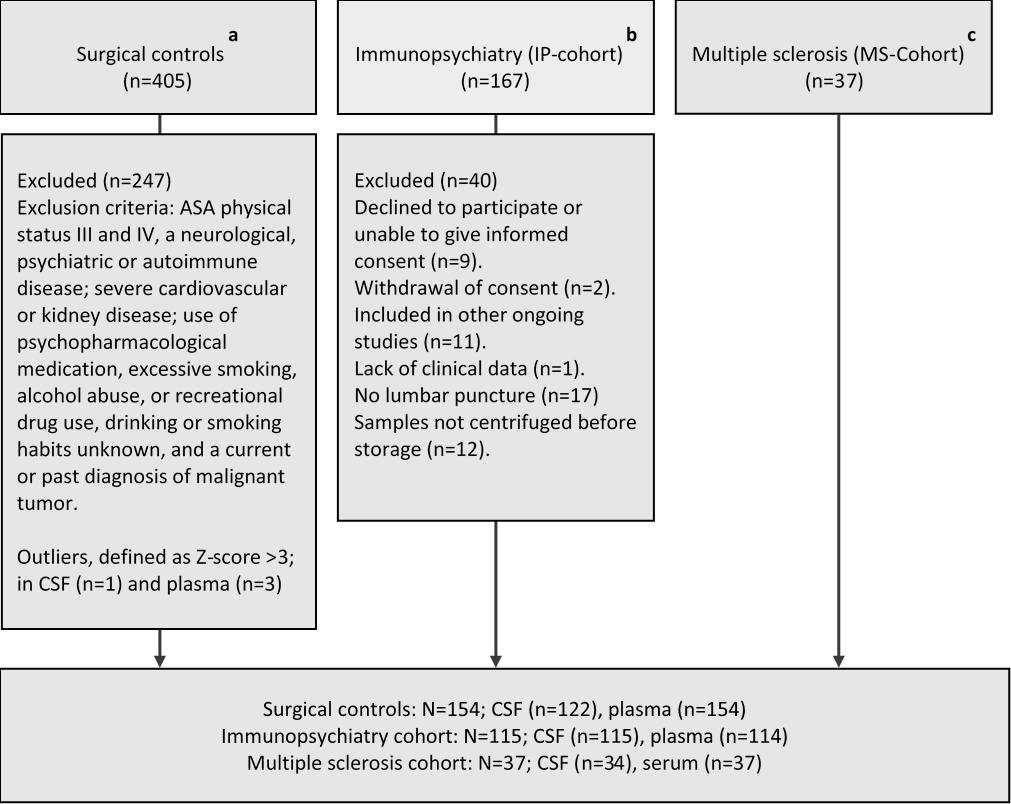

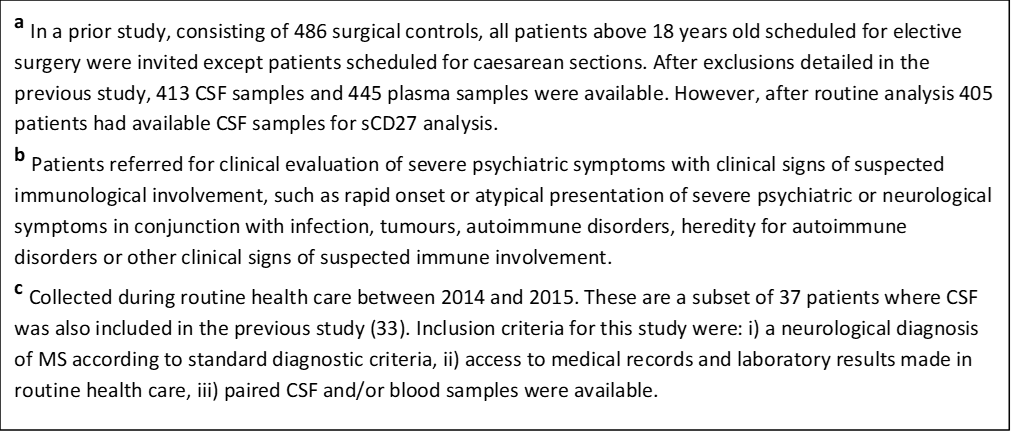

**Supplementary Figure 2**

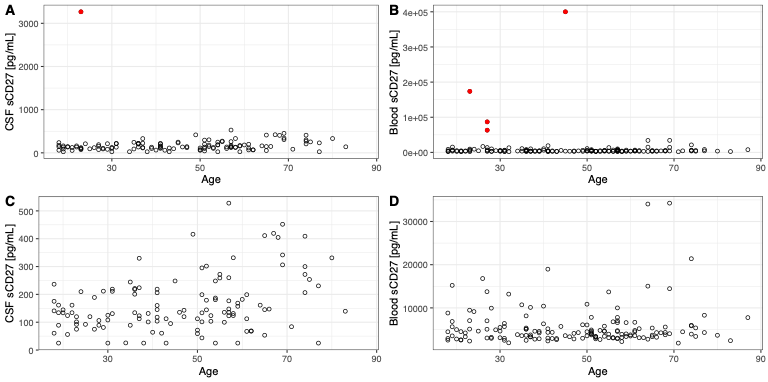
**Supplementary Figure 2.** **A correlation plot clearly demonstrating outliers for sCD27 levels in CSF and blood in healthy controls.** The outliers, defined as Z-score ˃3, are highlighted in red, n =4 in total. A-B) Controls sCD27 levels in CSF and blood before removal of outliers. C-D) Demonstration of controls’ sCD27 levels and age distribution after removal of outliers.

**Supplementary Figure 3.**

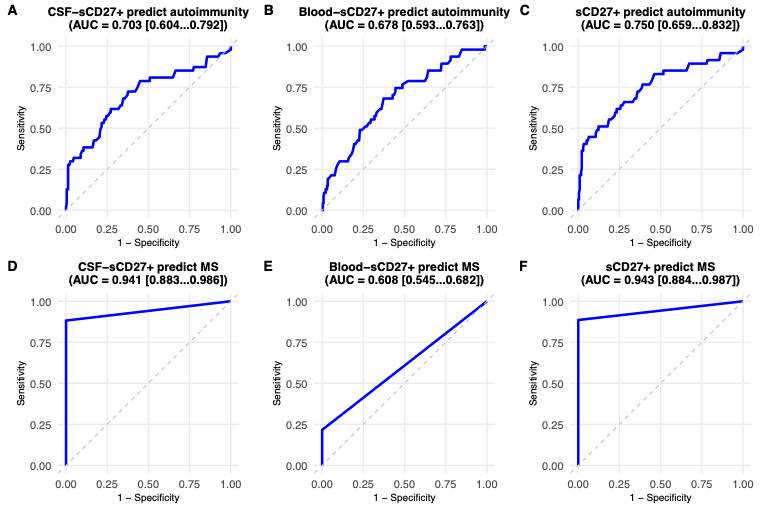

**Supplementary Figure 3.** **Receiver Operating Characteristic (ROC) curves showing that the combination of blood and CSF sCD27+ provides the highest predictive performance for comorbid autoimmune disease in patients with severe psychiatric disorders (IP-cohort) while CSF sCD27+ alone highly predicts multiple sclerosis (MS).** The plots evaluate logistic regression models distinguishing individuals with and without autoimmunity using: A) binary CSF-sCD27+ IP-cohort cases and healthy controls (AUC = 0.704 [0.605–0.790]); B) binary blood-sCD27+ IP-cohort cases and healthy controls (AUC = 0.677 [0.593–0.759]); C) binary blood or CSF-sCD27+ IP-cohort cases and healthy controls (AUC = 0.751 [0.662–0.832]); D) binary CSF-sCD27+ MS cohort cases and healthy controls (AUC = 0.941 [0.881–0.986]); E) binary blood-sCD27+ MS cohort cases and controls (AUC = 0.608 [0.543–0.683]); and F) binary blood or CSF-sCD27+ MS cohort cases and controls (AUC = 0.943 [0.882–0.987]). The diagonal dashed line indicates the line of no discrimination (AUC = 0.5).

**Supplementary Figure 4.**
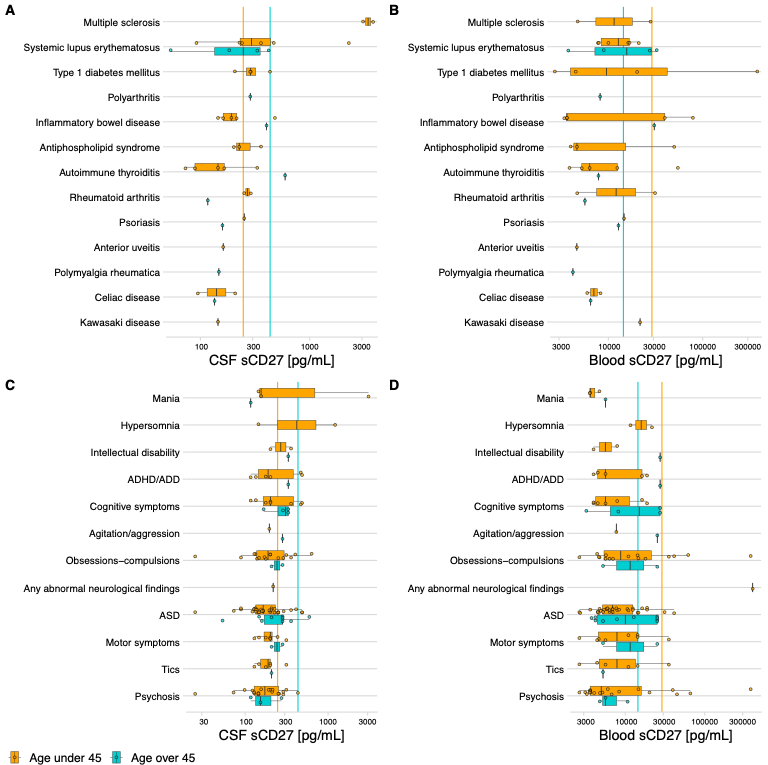
**Supplementary Figure 4. Distribution of sCD27 levels across autoimmune disease categories in patients with severe psychiatric disorders in the IP cohort.** Boxplots display log-transformed sCD27 levels with original labels in CSF and plasma. A-B) Distinct disease-specific patterns are observed across the two compartments. Elevated CSF sCD27 levels are most prominent in patients with comorbid Multiple sclerosis, systemic lupus erythematosus and type 1 diabetes (DM1) and Inflammatory bowel disease is more often elevated in plasma. These differences suggest compartmentalized immune activation that may reflect the primary site of immune involvement across autoimmune conditions. C-D) While headache is associated with elevated sCD27 in both compartments, other symptoms display distinct ranking patterns between CSF and plasma, suggesting compartment-specific immune associations. Inflammatory bowel disease included ulcerative proctitis, ulcerative colitis, Crohn’s disease, and microscopic colitis; type 1 diabetes mellitus included type 1 diabetes and type 1 diabetes/latent autoimmune diabetes in adults; rheumatoid arthritis included juvenile rheumatoid arthritis and rheumatoid arthritis; multiple sclerosis included multiple sclerosis and one case also has anti-N-methyl-D-aspartate receptor encephalitis; systemic lupus erythematosus included systemic lupus erythematosus and overlap diagnoses such as mixed connective tissue disease/systemic lupus erythematosus, systemic lupus erythematosus/myositis, and autoimmune hepatitis/systemic lupus erythematosus; and autoimmune thyroiditis included Hashimoto’s thyroiditis.

**Supplementary Figure 5.**

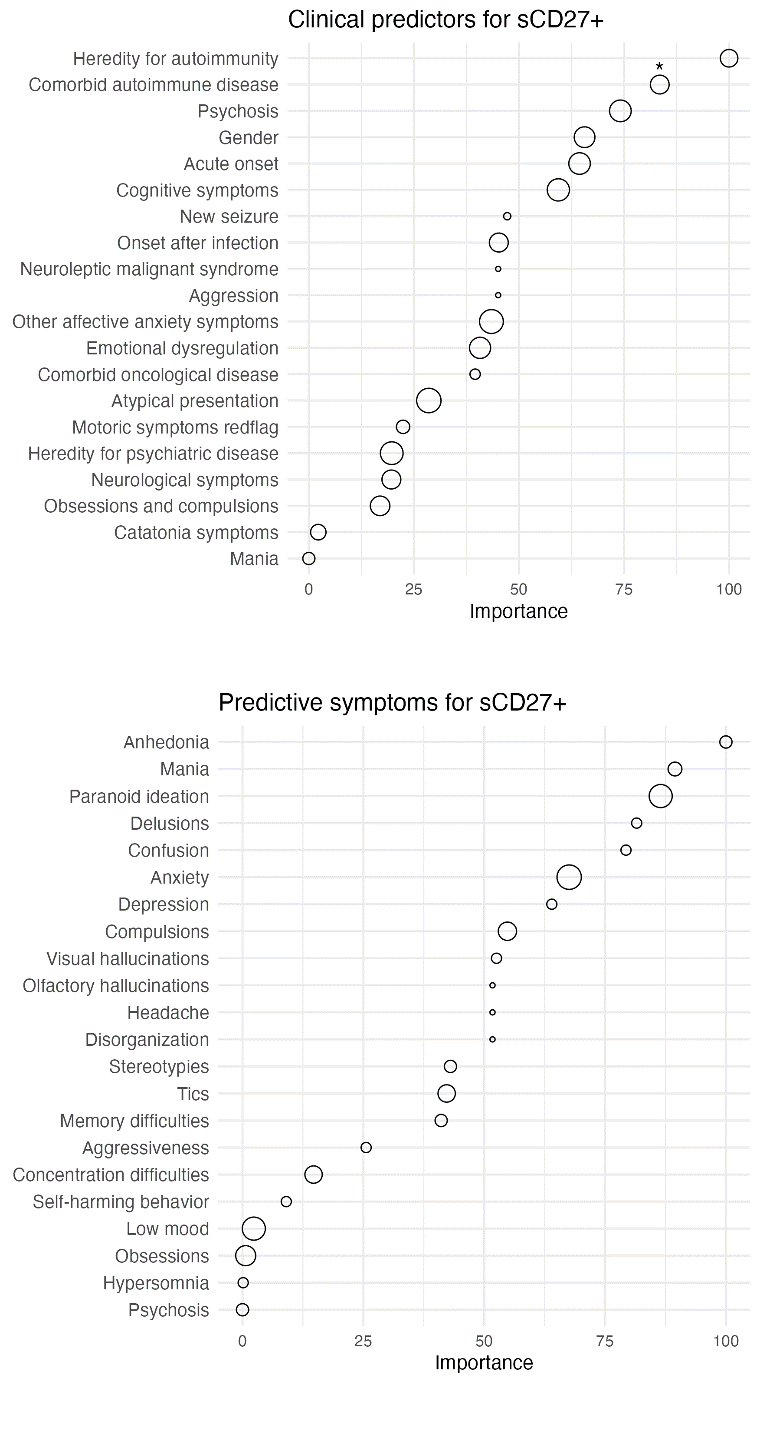

**Supplementary Figure 5. Feature importance plots from random forest models identifying clinical predictors of sCD27+ in both CSF and/or blood.** Clinical data and red flags for autoimmunity in psychiatry, top-ranking variables include cognitive symptoms, confusion and atypical presentation and psychosis.

**Supplementary Figure 6.**

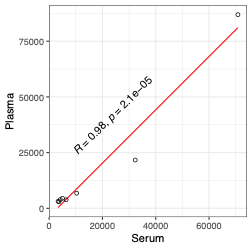

**Supplementary Figure 6. Relationship between concentrations measured in serum and plasma.** Individual measurements are shown as points, and the red line represents a linear regression fit without confidence intervals. A strong positive correlation was observed between serum and plasma values (Pearson’s r = 0.98, p = 2.1 × 10⁻⁵).

**Supplementary Table 1. Clinical Red flags in IP cohort (n=115)**

| Neuroleptic Malignant syndrome | Noted in the medical records | 2 |
| --- | --- | --- |
| Acute onset | (Sub)acute onset of severe psychiatric symptoms defined as a rapid progression of <3 months | 67 |
| Comorbid Oncological disease | Malignant or benign | 8 |
| Comorbid Autoimmune disorder | For example, systemic lupus erythematosus | 43 |
| Motor symptoms | For example, involuntary movements or dystonia | 19 |
| Neurological symptoms | Any abnormal findings in the neurological examination | 49 |
| Atypical presentation | A broad category including a global impression of atypical trajectory or symptomatology such as disproportionate cognitive deficits or hypersomnia | 95 |
| New seizures | Novel seizures in conjunction with or after disease onset | 3 |
| Catatonia symptoms | Catatonia symptoms reported by the assessing or treating clinician in the medical records or a total Busch Francis Catatonia Rating Scale score of >3 points | 31 |
| Cognitive symptoms | new memory loss, executive dysfunction, or disorientation | 74 |
| Prodromal infection | A temporal association of onset/deterioration of psychiatric/neurological symptoms with suspected or confirmed infection | 49 |

**Supplementary Table 2. Reference intervals for clinical markers** ^(41)^

| **NfL** | | **GFAp** | | **Total Tau** | | | |
| --- | --- | --- | --- | --- | --- | --- | --- |
| **Age (in years)** | **Reference value (ng/L)** | **Age (in years)** | **Reference value (ng/L)** | ***2013 to November 2019**** **Age (in years)** \| **Reference value (ng/L)** | | ***November 2019 to January 2020**** **Age (in years)** \| **Reference value (ng/L)** | |
| <30 | <380 | <20 | <175 | <18 | <250 | <18 | <300 |
| 30-39 | <560 | 20-59 | <750 | 18-44 | <300 | 18-49 | <360 |
| 40-59 | <890 | ≥60 | <1,250 | ≥45 | <400 | ≥50 | <409 |
| ≥60 | <1,850 |  | | | | | |
| *** The reference values were changed in November 2019  **IgG index:** (ref < 0.63)  **Q-Alb**: Age-related CSF/plasma albumin quotient (ref: age > 6 months to 45 years, < 6.8; age ≥ 45 years, < 10.2) **CSF cells:** White blood cell count (ref < 5 cells per 10^6^/L). | | | | | | | |
